# A Systematic Review and Meta-analysis of D-Dimer Levels in Patients Hospitalized with Coronavirus Disease 2019 (COVID-19)

**DOI:** 10.1101/2020.06.24.20139600

**Authors:** Agam Bansal, Achintya D Singh, Vardhmaan Jain, Manik Aggarwal, Samiksha Gupta, Rana P Padappayil, Mohak Gupta, Agrima Mian

## Abstract

**Aim:** To determine if the d-dimer levels are elevated in individuals with COVID 19 having worse clinical outcomes including all-cause mortality, ICU admission or ARDS

**Methods:** We conducted a systematic review and meta-analysis of published literature in Pubmed, Embase and Cochrane database through April 9, 2020 for studies evaluating the d-dimer levels in patients with and without a worse clinical outcome (all-cause mortality, ICU admission and ARDS). A total of 6 studies included in the meta-analysis.

**Results:** The values of d-dimer were found to be significantly increased in patients with the composite clinical end point than in those without (SMD, 1.67 ug/ml (95% CI, 0.72-2.62 ug/ml). The SMD of the studies (Tang et al, Zhou et al, Chen et al), which used only mortality as an outcome measure was 2.5 ug/mL (95% CI, 0.62-4.41).

**Conclusion:** The results of this concise meta-analysis suggest that d-dimer is significantly increased in patients having a worse clinical outcome (all-cause mortality, ICU admission or ARDS).

## Introduction

The 2019 novel coronavirus (2019-nCoV) or the severe acute respiratory syndrome corona virus 2 (SARS-CoV-2) first identified in Wuhan district in China has spread rapidly to more than 177 countries and was declared as a global pandemic on March 11^th^, 2020 (1). As of March 28^th^, there were 640,589 confirmed cases and 29,848 deaths globally (2). In up to 5% of infected patients, the disease may progress to critical form manifesting as hypoxic respiratory failure, multi organ dysfunction or shock and around 2.5% patients die from the infection (3). Laboratory predictors of clinical deterioration can aid in escalating the care of the patients with this infection and assist in appropriate triaging and resource utilization. Studies have reported an association of D-dimer >1 ug/ml with increased mortality in patients with COVID 19 infection (4). We systematically reviewed the current scientific literature to understand whether the measurement of D-dimers is associated with increased risk of having ICU admission, ARDS (acute respiratory distress syndrome) or all-cause mortality in patients hospitalized with COVID 19.

## Methods

### Literature Search

We carried out an electronic search in Medline (PubMed), Embase, and Cochrane database using the keywords “D-dimer” AND “Coronavirus 2019” OR “COVID 19” OR “SARS-CoV-2” OR “2019-nCoV”, between 2019 and current date (9^th^ April, 2020). Articles were limited to English language publications.

### Selection of studies

We applied the Preferred Reporting Items for Systematic Reviews and Meta-Analyses statement (PRISMA) to the methods for this study (5) (Figure 1). After duplications were removed, the title and abstracts were independently screened by two reviewers (AB and VJ). The studies reporting mean or the median D-dimer values in COVID 19 patients with and without a composite end point (ICU admission, ARDS and all-cause mortality) were included in the study. We excluded case reports, studies involving pediatric patient population and those not reporting the above-mentioned composite end points. We cross-referenced the research papers to identify additional studies meeting the inclusion criteria. Full texts of the included studies were then reviewed by two independent reviewers (AB and VJ) and data was extracted. Any conflicts were settled by a third author (ADS).

**Figure 1:**
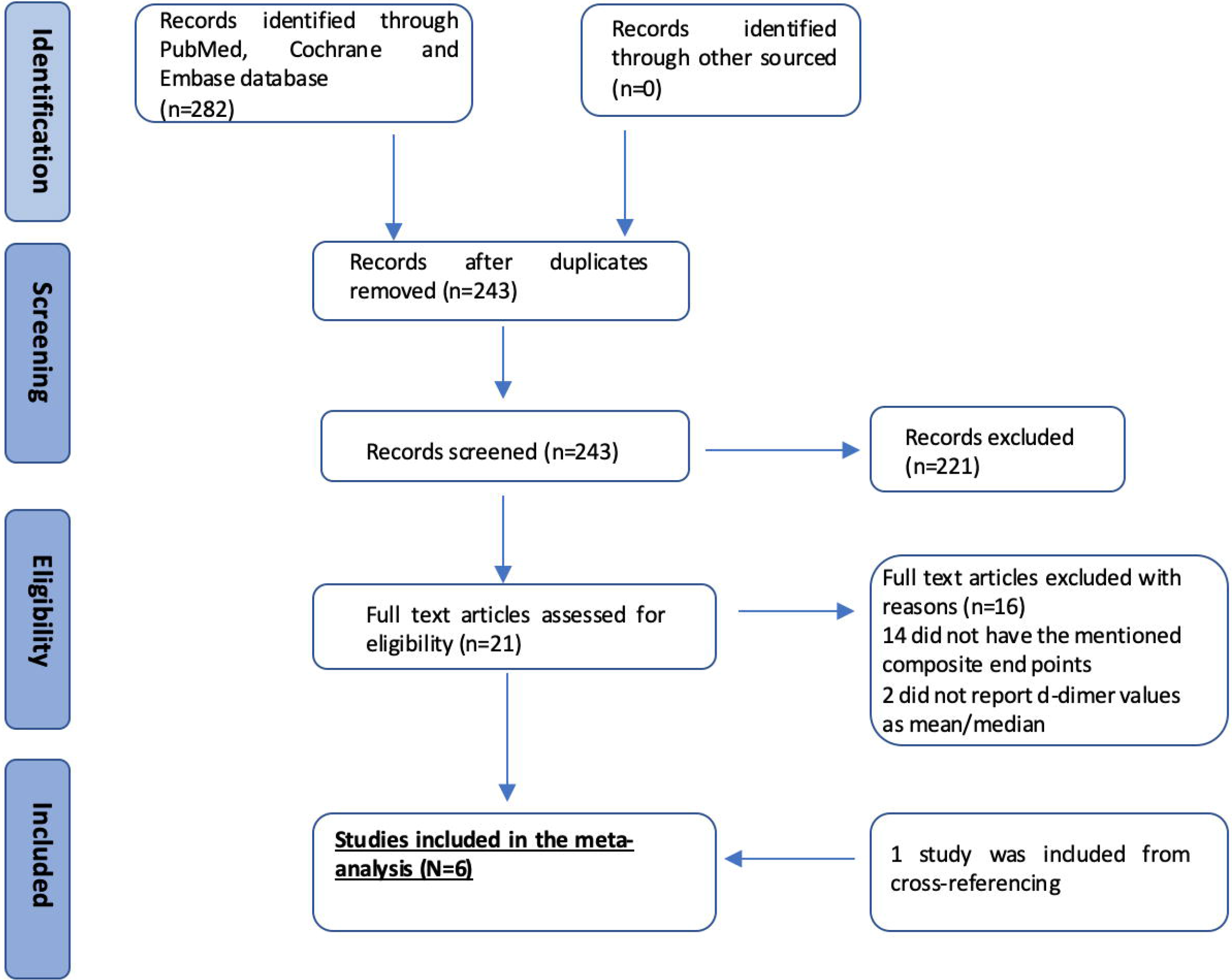
Preferred Reporting Items for Systematic Reviews and Meta-Analyses statement (PRISMA) flow chart for this study

**Figure 2:**
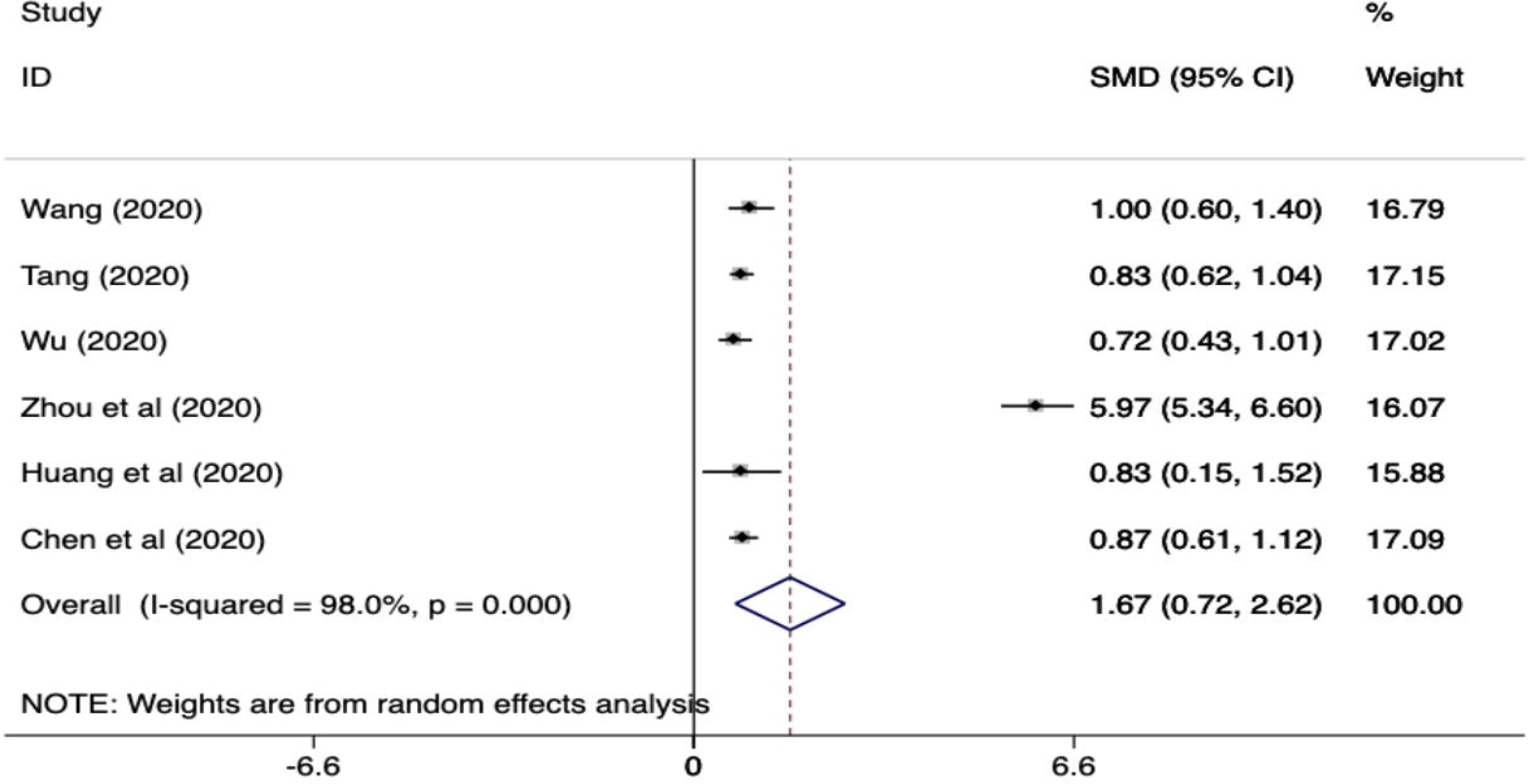
Standardized mean difference (SMD) and 95% confidence interval (CI) for predicting composite clinical end point (ARDS, ICU admission and mortality) in patients with COVID 19 infection

### Data extraction and Study Quality Appraisal

The following data variables were collected: author name, journal of publication, year published, country where the study was performed, type of study, number of patients, composite end point definition, and mean d-dimer values in patients with and without outcome of interest (all-cause mortality, ICU admission and ARDS).

Two authors (AB and VJ) independently assessed the risk of bias in the included studies using the validated Newcastle-Ottawa Scale.

### Statistical analysis

The meta-analysis was conducted with the calculation of standardized mean difference (SMD) and 95% confidence interval (95% CI) of D-dimer values in coronavirus 2019 patients with and without a composite clinical end point. D-dimer values were entered as a continuous variable. The mean and the standard deviation were extrapolated from the sample size, median and interquartile range (Q1-Q3) as per Hozo et al (6). I^2^ statistic was used to assess the heterogeneity between studies with values 0–30%, more than 30–60%, and more than 60% corresponding to low, moderate, and high degree of heterogeneity, respectively. DerSimonian and Laird random effects model was used for pooling the studies. The statistical analysis was performed using Stata 12 software (Stata Corp, College Station, Texas).

## Results

Our systematic electronic search resulted in 21 publications after the initial screening of titles and abstracts. Subsequently, 16 studies were excluded, yielding 5 studies that met the inclusion criteria for systematic review. Cross-referencing of full-text articles resulted in 1 additional study. Therefore, 6 studies were included in the final meta-analysis for association of mean/median d-dimer values with ICU admission, ARDS or mortality. Table 1 elucidates the baseline characteristics and outcomes of the included studies.

**Table 1:** Characteristics of the studies (n=6) included in the meta-analysis

| **Study** | **Zhou et al** | **Chen et al** | **Tang et al** | **Wang et al** | **Huang et al** | **Wu et al** |
| --- | --- | --- | --- | --- | --- | --- |
| **Study year** | 2020 | 2020 | 2020 | 2020 | 2020 | 2020 |
| **Study location** | Wuhan, China | Wuhan, China | Wuhan, China | Wuhan, China | Wuhan, China | Wuhan, China |
| **Study type** | Retrospective cohort | Retrospective cohort | Cross-sectional study | Retrospective cohort | Prospective Cohort | Retrospective Cohort |
| **Sample size** | 191 (Cases 54, Controls 137) | 274 (Cases 113, Controls 161) | 183 (Cases 21, Controls 162) | 138 (Cases 36, controls 102) | 41 (Cases 13, Controls 28) | 201 (Cases 84, Controls 117) |
| **Median age** | 56 (46-67) | 62 (44-70) | 54 (44-62) | 56 (42-68) | 49 (41-58) | 51 (43-60) |
| **Female** | 72 (38%) | 103(38%) | 85 (46.44%) | 63 (45.7%) | 11 (27%) | 73 (36.3%) |

| **Composite end point** | All-cause mortality (in-hospital) | All-cause mortality (in-hospital) | All-cause mortality (in-hospital) | ICU admission | ICU admission | ARDS (WHO definition) |
| --- | --- | --- | --- | --- | --- | --- |
| **Median D-dimer level, case and control** | Cases- 5.2 (1.5-21.1)<br>Controls- 0.6 (0.3-1.0) | Cases- 4.6 (1.3-21.0)<br>Controls- 0.6 (0.3-1.3) | Cases- 2.12 (0.77-5.27)<br>Controls- 0.61 (0.35-1.29) | Cases- 4.14 (1.91-13.24)<br>Controls- 1.66 (1.01-2.85) | Cases- 2.4 (0.6-14.4)<br>Controls- 0.5 (0.3-0.8) | Cases- 1.16 (0.46-5.37)<br>Controls- 0.52 (0.21-0.94) |

There were a total of 1329 patients with 434 (32.65%) patients having a composite clinical end point. The composite end point was defined as defined as mortality in 3 studies (4, 9, 11), ICU admission in 2 studies (8, 12) and onset of ARDS in another study (10). Zhou et al (4) showed the clinical and laboratory data of 191 hospitalized patients and observed that d-dimer levels were about 8-9 times higher in patients who died (median d-dimer 5.2 ug/ml, IQR:1.5-21.1 ug/ml) than those who survived (median d-dimer 0.6 ug/ml, IQR 0.3-1.0 ug/ml). Similarly, Chen et al (11) also observed an approximate seven-fold increase in d-dimer values in patients who had in-hospital all-cause mortality (median 4.6 ug/ml, IQR: 1.3-21.0 ug/ml) compared to patients who did not have the outcome (median 0.6 ug/ml, IQR: 0.3-1.3 ug/ml). Tang et al (9) showed a 3-4 times greater levels of d-dimer levels in patients who had in-hospital mortality compared to those who did not. Wang et al and Huang et al (8, 12) showed that d-dimers were significantly elevated in patients who required ICU admission. Furthermore, d-dimers were also significantly higher in patients having ARDS during the admission than those not having the outcome (10).

The standardized mean difference (SMD) for the six studies is summarized in Figure 1. The values of D-dimer were found to be significantly increased in patients with the composite clinical end point than in those without (SMD, 1.67 ug/ml (95% CI, 0.72-2.62 ug/ml). The SMD of the studies (Tang et al (9), Zhou et al (4), Chen et al (11)), which used only mortality as an outcome measure was 2.5 ug/mL (95% CI, 0.62-4.41). The heterogeneity of the studies was found to be relatively high (i.e. I^2^ statistic 98%).

There were two additional studies which reported higher d-dimer levels in patients with worse outcomes. However, they were not included in our meta-analysis as they did not report the median/mean d-dimer levels. Zhang et al (13) described the characteristics of 95 patients and found that out of the 25 patients having an outcome (ICU admission, mechanical ventilation or death), 23 (92%) had d-dimer values ≥ 1 ug/ml. Similarly, another study (14) showed around 70% of the patients with worse outcome (ICU admission, mechanical ventilation or death) having d-dimers ≥ 0.5 ug/ml.

## Discussion

In this study, we performed a systematic review and meta-analysis of studies to assess whether the d-dimer levels were associated with a composite end points including all-cause mortality, ICU admission and ARDS in patients hospitalized with COVID 19. We found that 1) d-dimers were significantly elevated in patients having a composite end point compared to those not having the outcome, 2) the level of d-dimers was higher in studies having mortality as an outcome in comparison to other end-points.

The normal value of d-dimer is <0.5 ug/ml. There are several plausible reasons for elevated D-dimer in patients hospitalized with COVID 19 having worse clinical outcomes. First, patients with severe COVID 19 infection can have DIC (disseminated intravascular coagulation) secondary to sepsis. Severe acute lung injury or ARDS by itself has also been associated with increased incidence of DIC. Tang et al (9) mentioned in their study that the vast majority of patients who died during admission fulfilled the criteria for DIC (71.6% vs 0.6% in survivors). Second, prior studies have shown that severe acute respiratory infection can cause injury to the endothelial cells and increase the levels of hemostatic factors such as d-dimers and vWF (15). Third, respiratory infections have been associated with deep vein thrombosis and pulmonary embolism. Wang et al (16) postulated about the possible formation of pulmonary microthrombus in patients infected with H1N1 infection and a consequent elevation in D-dimer. There have been 2 cases reported of pulmonary embolism in COVID 19 infected patients (17). Fourth, the COVID-19 patients with critical form of the disease are more likely to have additional complications including acute kidney injury, acute cardiac injury, congestive heart failure, all of which can cause increase the levels of D-dimers. Finally, the elderly patients are at an increased risk of having worse clinical outcomes from COVID 19 infection and d-dimers are higher in elderly patient population.

The major limitation of the studies included was that it is unknown as to when were the d-dimer levels obtained during the course of admission. In addition, there was a significant heterogeneity in the reported results. This was likely due to differences in study size, selection bias, and different stages at which the D-dimer values were measured. Also, since all the studies included have been performed in China, the external validity is lacking.

The results of this concise meta-analysis suggest that d-dimer is significantly increased in patients having a worse clinical outcome (all-cause mortality, ICU admission or ARDS). Further studies are required to assess if the serial measurement of d-dimer plays any role in predicting evolution towards a more critical form of disease. Also, it will be imperative to know if anticoagulation therapies are of use in patients with severe COVID 19 disease.

## Data Availability

This is a meta-analysis with all the data available

